# Immune transcriptomes from hospitalized patients infected with the SARS-CoV-2 variants B.1.1.7 and B.1.1.7 carrying the E484K escape mutation

**DOI:** 10.1101/2021.05.27.21257952

**Authors:** Hye Kyung Lee, Ludwig Knabl, Ludwig Knabl, Manuel Wieser, Anna Mur, August Zabernigg, Jana Schumacher, Norbert Kaiser, Priscilla A. Furth, Lothar Hennighausen

**Affiliations:** National Institute of Diabetes, Digestive and Kidney Diseases, Bethesda, MD 20892, USA; TyrolPath, Zams, Austria; Krankenhaus St. Vinzenz, Zams, Austria; Division of Internal Medicine, Krankenhaus Kufstein, Kufstein, Austria; Division of Internal Medicine, Krankenhaus St. Johann, St. Johann, Austria; Departments of Oncology & Medicine, Georgetown University, Washington, DC, USA

## Abstract

Fast-spreading variants of severe acute respiratory syndrome coronavirus 2 (SARS-CoV-2) energize the COVID-19 pandemic. B.1.1.7 (VOC-202012/01) has become the predominant variant in many countries and a new lineage (VOC-202102/02) harboring the E484K escape mutation in the B.1.1.7 background emerged in February 2021^1^. This variant is of concern due to reduced neutralizing activity by vaccine-elicited antibodies^2,3^. However, it is not known whether this single amino acid change leads to an altered immune response. Here, we investigate differences in the immune transcriptome in hospitalized patients infected with either B.1.1.7 (n=28) or B.1.1.7+E484K (n=12). RNA-seq conducted on PBMCs isolated within five days after the onset of COVID symptoms demonstrated elevated activation of specific immune pathways, including JAK-STAT signaling, in B.1.1.7+E484K patients as compared to B.1.1.7. Longitudinal transcriptome studies demonstrated a delayed dampening of interferon-activated pathways in B.1.1.7+E484K patients. Prior vaccination with BNT162b vaccine (n=8 one dose; n=1 two doses) reduced the transcriptome inflammatory response to B.1.1.7+E484K infection relative to unvaccinated patients. Lastly, the immune transcriptome of patients infected with additional variants (B.1.258, B.1.1.163 and B.1.7.7) displayed a reduced activation compared to patients infected with B.1.1.7. Acquisition of the E484K substitution in the B.1.1.7 background elicits an altered immune response, which could impact disease progression.

## Introduction

Over the past few months, several variants of concern (VOC) carrying specific mutations thought to enhance viral fitness have emerged. Specifically, B.1.351^4^ and P.1 were of particular concern because they carry the mutation E484K within the receptor binding domain (RBD), which has been demonstrated to enhance escape from neutralizing antibody inhibition *in vitro*^2,3^ and may be linked with reduced vaccination efficacy^5^. In February 2021 Public Health England (PHE) published a report of B.1.1.7 genomes with acquisition of the E484K spike mutation^1^. Since the PHE announcement a total of 1332 B.1.1.7+E484K genomes have been uploaded to GISAID from England and 14 other countries, including 203 from the United States where it was first detected in Oregon (as of 05/24/2021).

The new B.1.1.7+E484K variant (VOC 202102/02) is fast spreading, prompting interest in the response of both unvaccinated and vaccinated individuals to disease from this variant. Recent *in vitro* data demonstrated that introduction of the E484K mutation into the B.1.1.7 background led to a more-substantial loss of neutralizing activity by vaccine-elicited antibodies compared with the mutations in B.1.1.7 alone^2,3^, suggesting that this variant represents a threat to the efficacy of the BNT162b2 vaccine. However, it remains unclear whether the B1.1.7+E484K variant elicits a modified immune response in patients compared with the parent B.1.1.7 variant.

To obtain insight into the biological significance of the E484K mutation in the B.1.1.7 backbone, we investigated the transcriptome of a total of 40 hospitalized patients infected with either the B.1.1.7+E484K variant (n=12) or the B.1.1.7 parent strain (n=28). We also investigated the impact of the BNT162b vaccine on both variants.

## Results

### Immune transcriptome progression after B.1.1.7 infection

Transcriptomes from 28 patients infected with the B.1.1.7 variant and 12 patients infected with the B.1.1.7+E484K variant were compared (Table 1). First, we investigated the temporal progression of the immune transcriptomes in 28 patients hospitalized with infection of the B.1.1.7 variant (Table 1, Fig. 1a, Supplementary Table 1). RNA-seq experiments were conducted on PBMCs isolated between days 1-5 (group A), days 10-14 (group B) and days 15-30 (group C) after reporting symptomology (Supplementary Data 1-4). The presence of SARS-CoV-2 was confirmed by PCR, followed by whole viral genome sequencing. PCA plots demonstrate a separation of the three cohorts, with group C shifting towards the non-COVID controls (Fig. 1b). Expression of a total of approximately 3,071 genes was significantly elevated within five days of reporting symptoms and these clustered in immune-relevant pathways (Fig. 1c). Expression of immune-related gene classes, including the highly activated JAK/STAT pathway, declined up to 95% after 15 days upon onset of COVID-19 (Fig. 1c and d, Supplementary Fig. 1).

**Table 1.** Demographic and Clinical Characteristics of COVID-19 patients infected by the B.1.1.7 or B.1.1.7+E484K SARS-CoV-2 variants.

| Type of SARS-CoV-2 virus | B.1.1.7/E484K | B.1.1.7 | Significant | Chi-square | *p* value |
| --- | --- | --- | --- | --- | --- |
| Patients, *n* (%) | 12 (30) | 28 (70) |  |  |  |
| Patient age (mean) | 70.30 | 72.40 | No | 0.41 | 0.94 |
| > 65yr, *n* (%) | 8 (66.7) | 20 (71.4) |  |  |  |
| 41-65, *n* (%) | 3 (25.0) | 7 (25.0) |  |  |  |
| 21-40, *n* (%) | 1 (8.3) | 1 (3.6) |  |  |  |
| Gender |  |  | No | 0.94 | 0.62 |
| females, *n* (%) | 4 (33.3) | 14 (50.0) |  |  |  |
| males, *n* (%) | 8 (66.7) | 14 (50.0) |  |  |  |
| BMI (mean) | 26.5 | 25 | No | 0.68 | 0.95 |
| underweight (<18.5), *n* (%) | 1 (8.3) | 1 (3.6) |  |  |  |
| normal (18.5-24.9), *n* (%) | 4 (33.3) | 12 (42.9) |  |  |  |
| overweight (25.0-29.9), *n* (%) | 4 (33.3) | 8 (28.6) |  |  |  |
| obese (≥30.0), *n* (%) | 1 (8.3) | 3 (10.7) | | | |
| Vaccination, *n* (%) | 3 (25.0) | 5 (17.9) | No | 0.17 | 0.68 |
| Treatment, *n* (%) | 7 (58.3) | 18 (64.3) | No | 0.85 | 0.84 |
| corticosteroid | 7 (58.3) | 17 (60.7) |  |  |  |
| convalescent plasma | 0 | 2 (7.1) |  |  |  |
| ICU, *n* (%) | 3 (25.0) | 4 (14.3) | No | 0.17 | 0.68 |
| Deaths, *n* (%) | 3 (25.0) | 5 (17.9) | No | 0.17 | 0.68 |
| Data set | 16 | 47 | No | 6.70 | 0.08 |
| group A, *n* (%) | 7 (43.8) | 14 (29.8) |  |  |  |
| group B, *n* (%) | 9 (56.3) | 18 (38.3) |  |  |  |
| group C, *n* (%) | 0 | 15 (31.9) |  |  |  |

**Figure 1.**
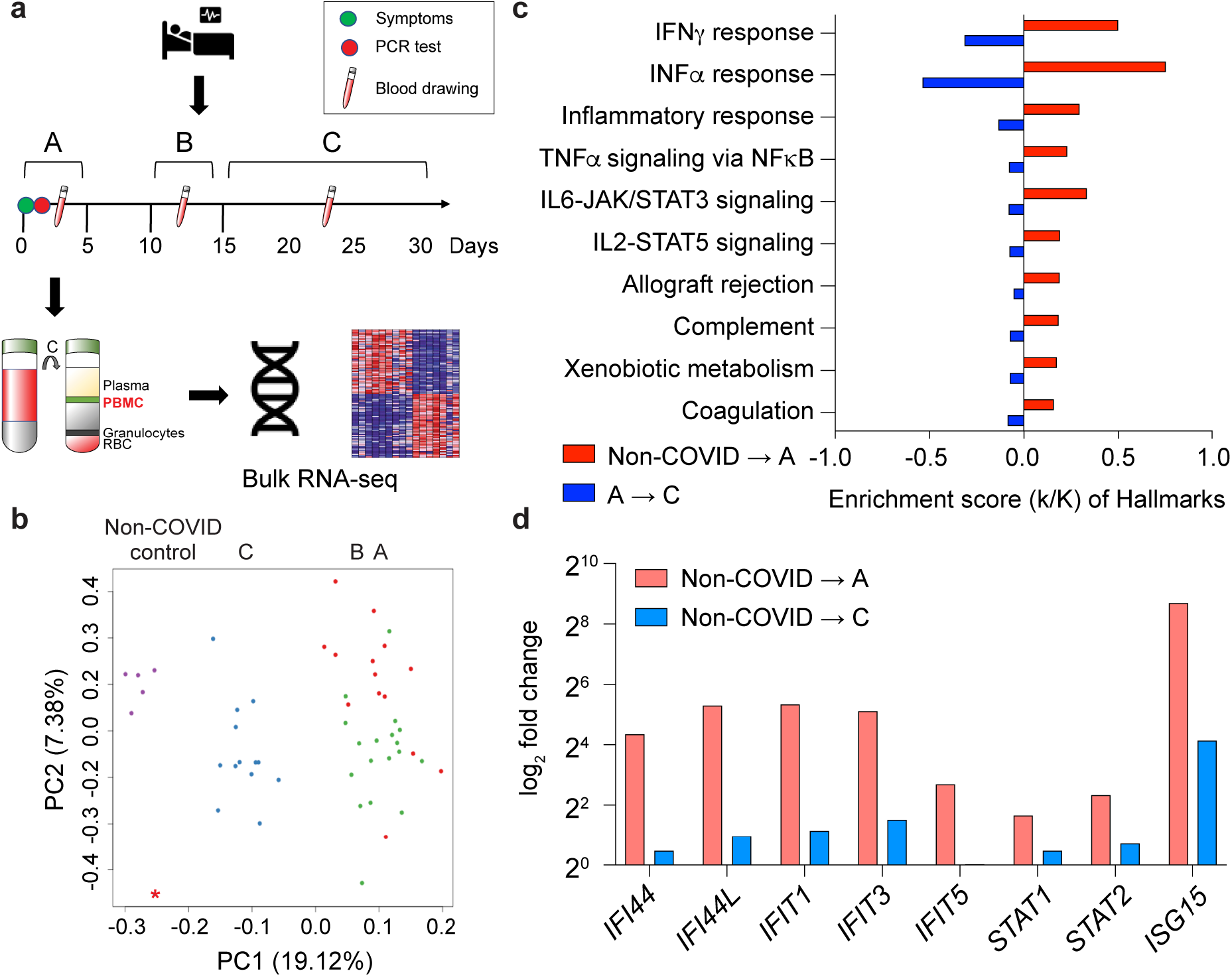
Temporal progression of immune transcriptomes in patients infected with B.1.1.7. **a**. Schematic presentation of the experimental workflow. COVID-19 patients were diagnosed, and the variants were identified through whole viral genome sequencing. PBMCs were purified from blood was drawn at different times after symptomology and RNA-seq was performed. **b**. Principal-component analysis (PCA) of transcriptomes of 26 and five non-COVID controls^6^, depicting the variation in the global gene expression profiles across the three time points and non-COVID controls. Principal components 1 (PC1) and 2 (PC2), which represent the greatest variation in gene expression, are shown. **c**. Expression of immune pathway genes induced between days 1-5 (A) from the symptom onset was mitigated after approximately 15 days of symptom onset (FDR q value < 0.005). **d**. representative genes whose expression is highly induced within the first five days of symptomology and rapidly declines after two weeks.

### Comparison of early response immune transcriptomes of patients infected with B.1.1.7+E484K to those of patients infected with B.1.1.7

Next, we investigated whether the E484K escape mutant in the backbone of B.1.1.7 (B.1.1.7+E484K) leads to an altered immune response. For this, we compared the immune transcriptomes of patients infected with either the B.1.1.7+E484K or the B.1.1.7 parent variant within the first five days after the onset of COVID-19 symptomology. All patients had had been hospitalized and there were no significant differences in age distribution, gender, BMI, vaccination status, treatment, ICU and deaths between the two groups (Table 1). RNA-seq was conducted on seven B.1.1.7+E484K patients and 14 B.1.1.7 patients (Fig. 2; Supplementary Data 2, 5, 6). PCA plots showed separation between RNA-seq samples from non-COVID controls^6^ and the two variants on first and second principal components (PC1 and PC2) (Fig. 2a). A higher number of differentially expressed genes (DEGs) were identified between patients infected with the B.1.1.7+E484K variant compared to controls (4399) than found in the comparison between patients infected with the B.1.1.7 variant and controls (3071) (Fig. 2b). Hallmark Gene Set Enrichment Analysis (GSEA) established that the significantly up-regulated genes in both cohorts were enriched in immune response pathways, including IL-JAK/STAT signaling and IFNα/γ responses (Fig. 2c). Pathway analyses identified genes that are induced in lung tissue from COVID-19 patients and in cells infected by SARS-CoV-2 (Fig. 2d). Genes preferentially activated in the B.1.1.7+E484K variant are part of the interferon and JAK/STAT pathways (Fig. 2d-e; Supplementary Fig. 2-4). These experiments demonstrate that the acquisition of the E484K escape mutation in the B.1.1.7 background elicits a significantly different immune response than the parent variant itself.

**Figure 2.**
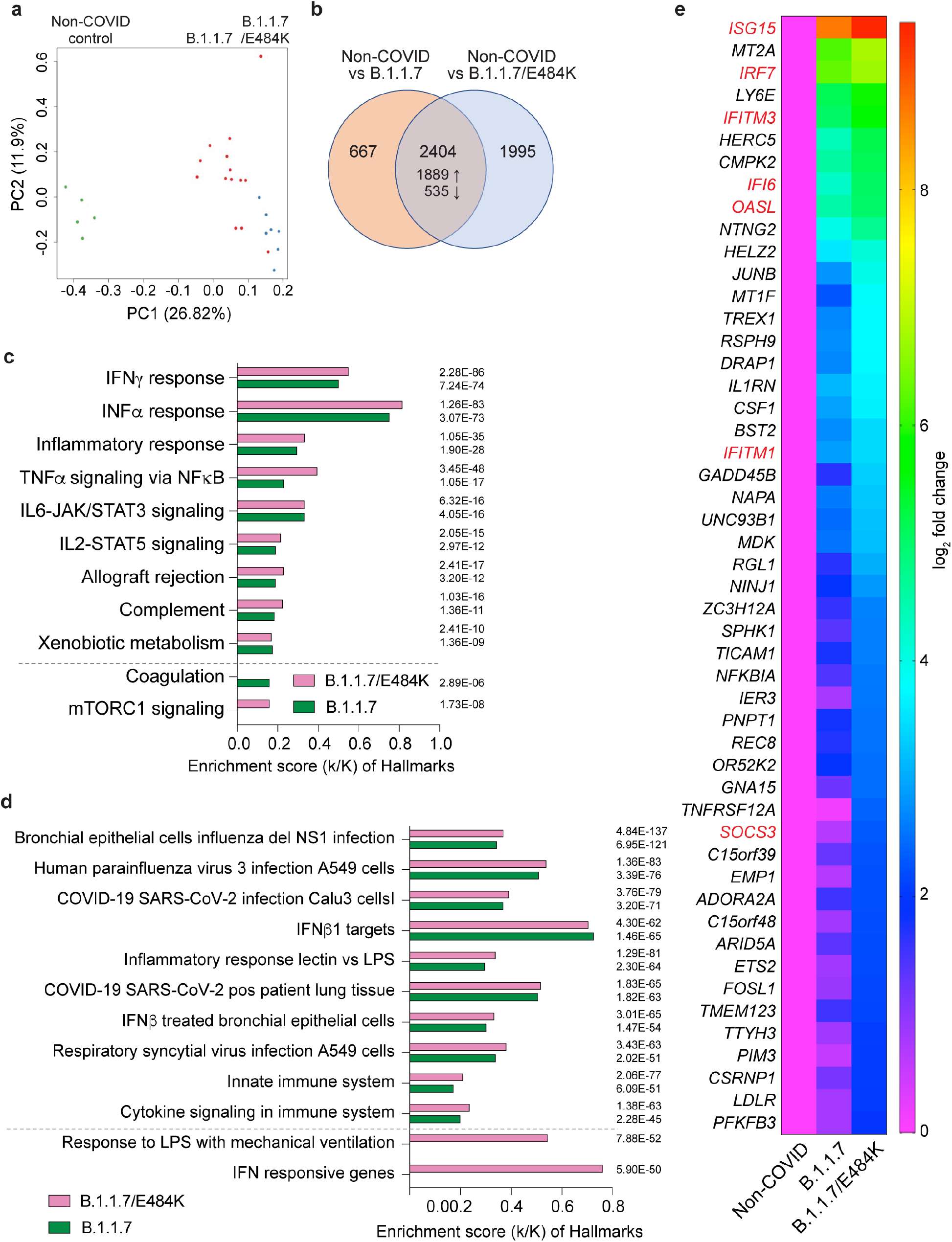
Differences in the immune transcriptome between patients infected with the B.1.1.7 or B.1.1.7+E484K variants and non-COVID controls. **a**. PCA plot of COVID-19 samples from the fourteen B.1.1.7 (red) and seven B.1.1.7+E484K (blue) patients based on the expression of all genes. Transcriptomes were established within the first five days of symptomology. **b**. More than one half of the genes whose expression was induced significantly in COVID-19 patients compared to non-COVID controls were shared between both patient groups. **c**. Genes expressed at significantly higher levels in the COVID-19 patients were significantly enriched in Hallmark Gene Sets (FDR q value < 0.005 and statistic of each hallmark is added next to bars). Nine hallmark sets involved in immune regulation were presented. **d**. Canonical pathway analysis of significantly enriched genes included in clusters for SARS-CoV-2 infection and innate immune-related biological processes. **e**. Comparison of relative normalized expression levels of IFI and ISG gene families are presented in box plot.

### Differences in temporal progression of the immune response in patients following B.1.1.7+E484K as compared to B.1.1.7 infection

Next, we utilized temporally matched samples from patients infected with B.1.1.7 (n=5) and B.1.1.7+E484K (n=4) to compare temporal progression of the immune transcriptome at days 1-5 versus 10-14 following symptomology (Supplementary Data 7-9). PCA plots demonstrated that while the transcriptomes from B.1.1.7 patients diverged on both PC1 and PC2 (Fig. 3a, left panel), the transcriptomes from B.1.1.7+E484K patients diverged only on PC2 (Fig. 3a, right panel). At days 1-5, 294 genes were preferentially expressed in B.1.1.7+E484K patients compared to B.1.1.7 with genes mapping to interferon and JAK/STAT pathways (Fig. 2e; Supplementary Fig. 3-4). The pronounced interferon and inflammatory responses seen at days 1-5 sharply declined between days 10-14 (Fig. 3b, c). Notably, the decrease was more pronounced in the B.1.1.7 variant with a more sustained immune response in the B.1.1.7+E484K variant. Since B.1.1.7+E484K induced a more expansive immune transcriptome than the B.1.1.7 parent variant, we directly compared their transcriptomes between days 10-14 after symptomology (Fig. 3d-e). The transcriptomes separate on the PCA plot (Fig. 3d) and the significantly higher expressed genes in B.1.1.7+E484K are enriched in cytokine responsive signaling and viral, including SARS-CoV-2, infection pathway (Fig. 3e-f). Of the 72 genes enriched in COVID-19 pathway (Fig. 3f), the top 50 induced gene in the B.1.1.7+E484K variant compared to B.1.1.7 at the second time point are related to interferon and JAK/STAT signaling (Supplementary Fig. 5).

**Figure 3.**
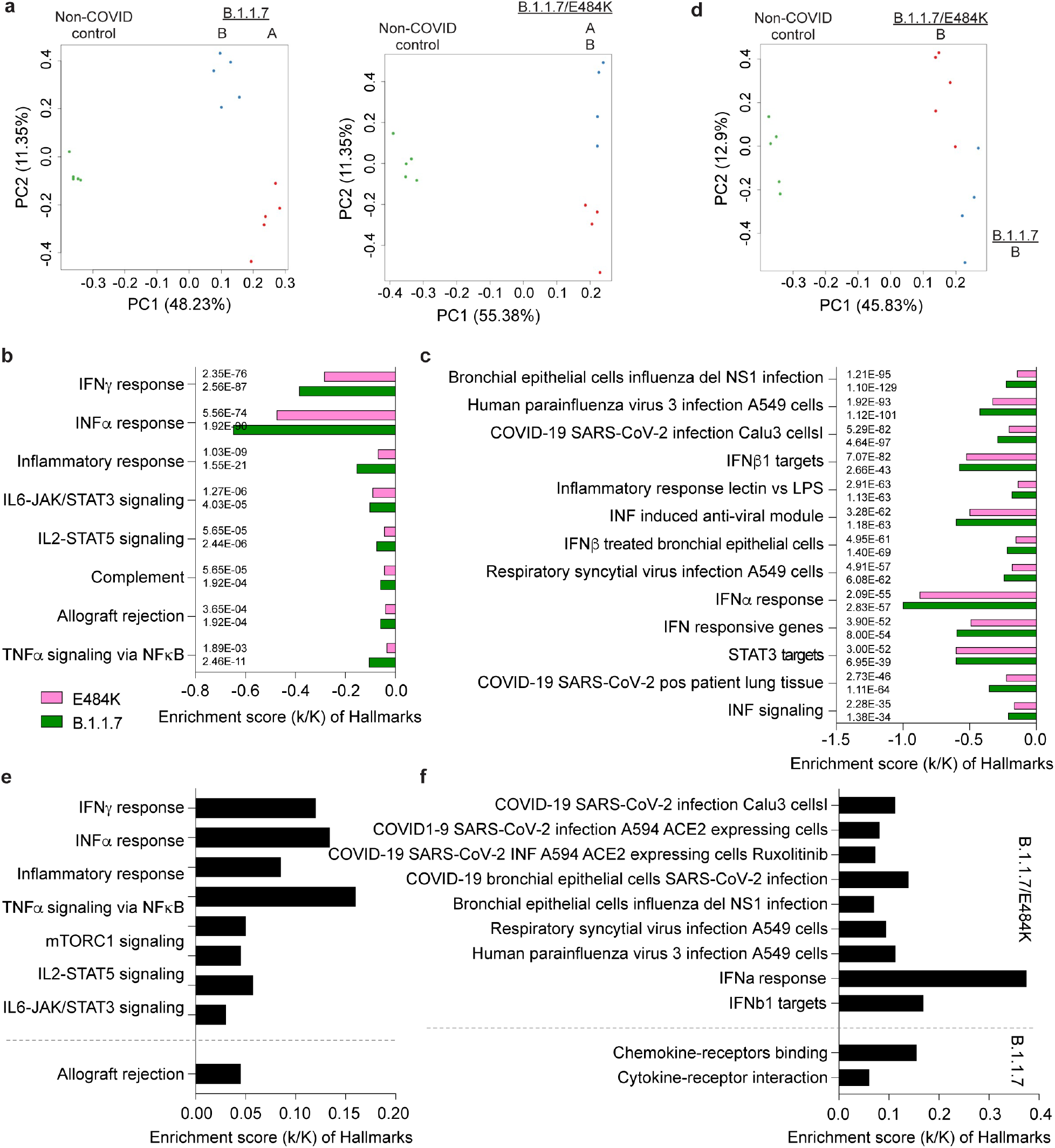
Longitudinal progression of immune transcriptomes from patients infected with the B.1.1.7 or B.1.1.7+E484K variants. **a**. Sparse PCA depicting global transcriptional profiles of samples from the same COVID-19 patients (five patients in B.1.1.7 variant and four patients in B.1.17/E484K variant) collected at days 1-5 (sample A in Fig. 1a) and 10-15 (sample B in Fig. 1a) after the onset of symptoms. **b-c**. Genes expressed at significantly higher levels in the COVID-19 patients for each variant were significantly enriched in Hallmark Gene Sets (b) and canonical pathway analysis (c) (FDR q value < 0.005 and statistic of each mark is added next to bars). Genes map to virus infection and immune regulation. **d**. PCA plot of non-COVID controls and transcriptomes from day 10-15 (sample B in Fig. 1a) of both COVID-19 patient groups (B.1.1.7 in red and B.1.1.7+E484K in blue) on the basis of the expression of all genes. **e-f**. Box plots showing hallmark and canonical pathway analysis enriched in DEGs. Most hallmarks of transcripts were included in innate immune-related biological processes. More than half of significant genes are enriched for anti-viral immune responses, including SARS-CoV-2 infection.

### Immune transcriptomes in vaccinated patients infected with B.1.1.7 and B.1.1.7+E484K

Next, we investigated the impact of the BNT162b vaccine on the immune transcriptome in the seven patients that had received at least one dose between 6 and 24 days prior to demonstrating COVID-19 symptomology and one fully vaccinated patient (Supplementary Data 10-12). Three vaccinated patients infected with B.1.1.7 died in the hospital (vaccination days 8, 10, 14 prior to symptoms). The transcriptomes from the three B.1.1.7+E484K patients who received their first dose between 8 and 22 days prior to developing symptoms were compared with six unvaccinated B.1.1.7+E484K patients (Fig. 4a). Expression of 19 genes was significantly reduced in the vaccinated cohort as compared to the unvaccinated one (Fig. 4b). These genes map to interferon, JAK/STAT and TNFα pathways (Fig. 4c-d). Differences were reflected in a significant difference on the PC2 but not PC1 component of the PCA plot (Supplementary Fig. 6). A direct comparison of the transcriptomes from three B1.1.7 and three B.1.1.7+E484K patients that had received at least one dose highlighted the different responses (Fig. 4e-g).

**Figure 4.**
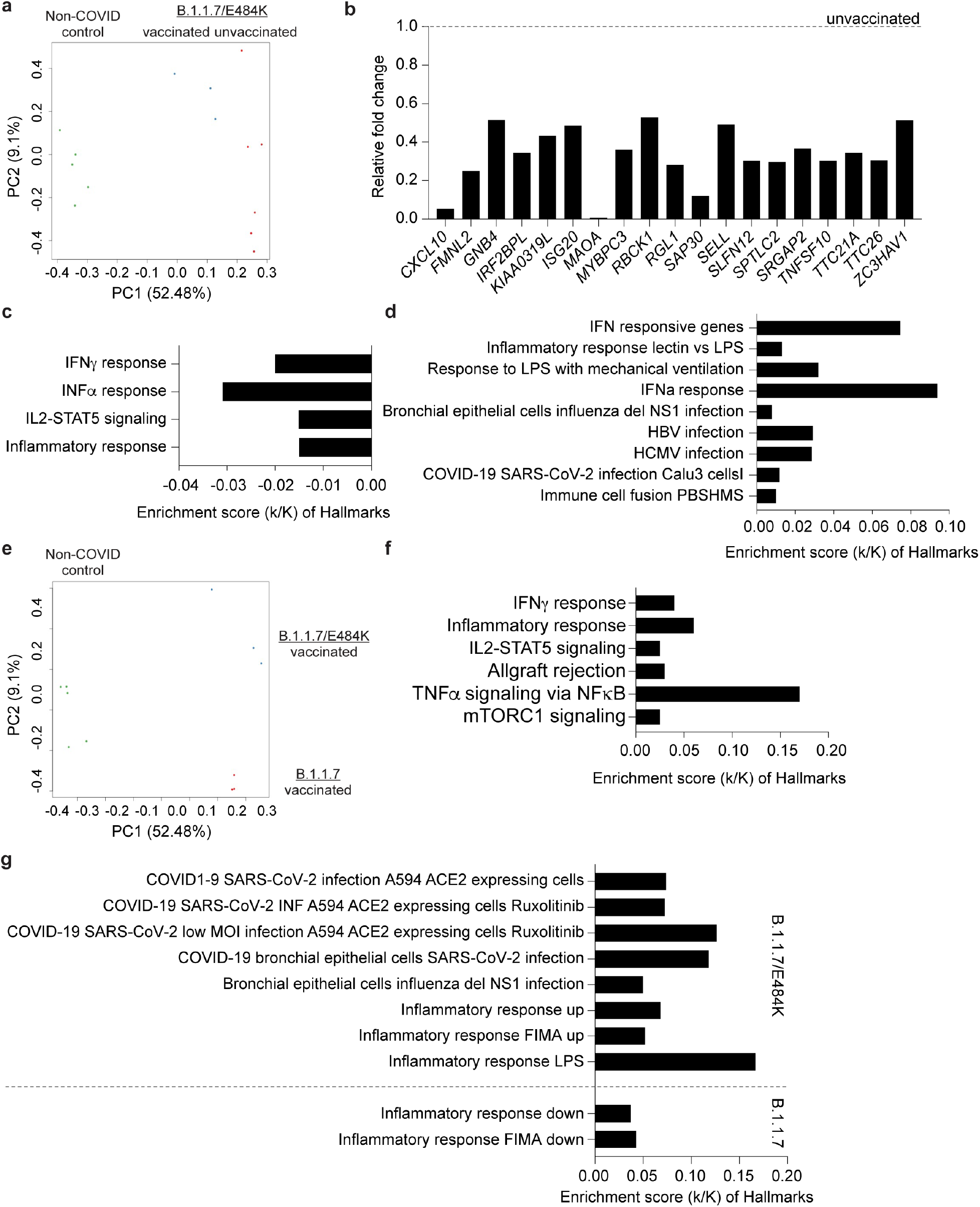
Differences of immune response between non-vaccinated and vaccinated COVID-19 patients. **a**. PCA plot of COVID-19 samples from the six unvaccinated (red) and three vaccinated (blue) COVID-19 patients infected by B.1.1.7+E484K on the basis of the expression of all genes. Samples are color-coded by vaccination status. **b**. Relative mRNA levels of 19 genes that are reduced in vaccinated patients as compared to unvaccinated ones are presented by bar graphs. **c-d**. Bar plots show the GSEA of genes expressed higher in vaccinated B.1.1.7+E484K patients than unvaccinated patients against Hallmark (b) and canonical pathway analysis (c) (FDR q value < 0.005). **e**. PCA plot of COVID-19 samples from the three vaccinated B.1.1.7 patients (red) and B.1.1.7/E484K patients (blue), respectively. **f-g**. The enrichment of 303 genes expressed higher in B.1.1.7+E484K patients than in B.1.1.7 patients is shown using bar plots. Hallmark (f) and canonical pathway analysis (g) (FDR q value < 0.005).

### Immune transcriptomes in patients infected with other SARS-CoV-2 variants

We also examined the immune transcriptome of patients infected with the variants B.1.177 (n=1), B.1.1.163 (n=2) and B.1.258 (=2) within the first five days of symptomology (Supplementary Table 1; Fig. 5a; Supplementary Data 13-16). COVID-19 patients are distributed by the type of variant on the PCA plot (Fig. 5b). The B.1.1.7 variant elicited stronger interferon responses as indicated by elevated JAK/STAT and inflammatory pathways compared to the other variants (Fig. 5c). Key components of ISGs are expressed highest in patients infected with B.1.1.7+E484K and B.1.1.7 compared to the other variants (Fig. 5d).

**Figure 5.**
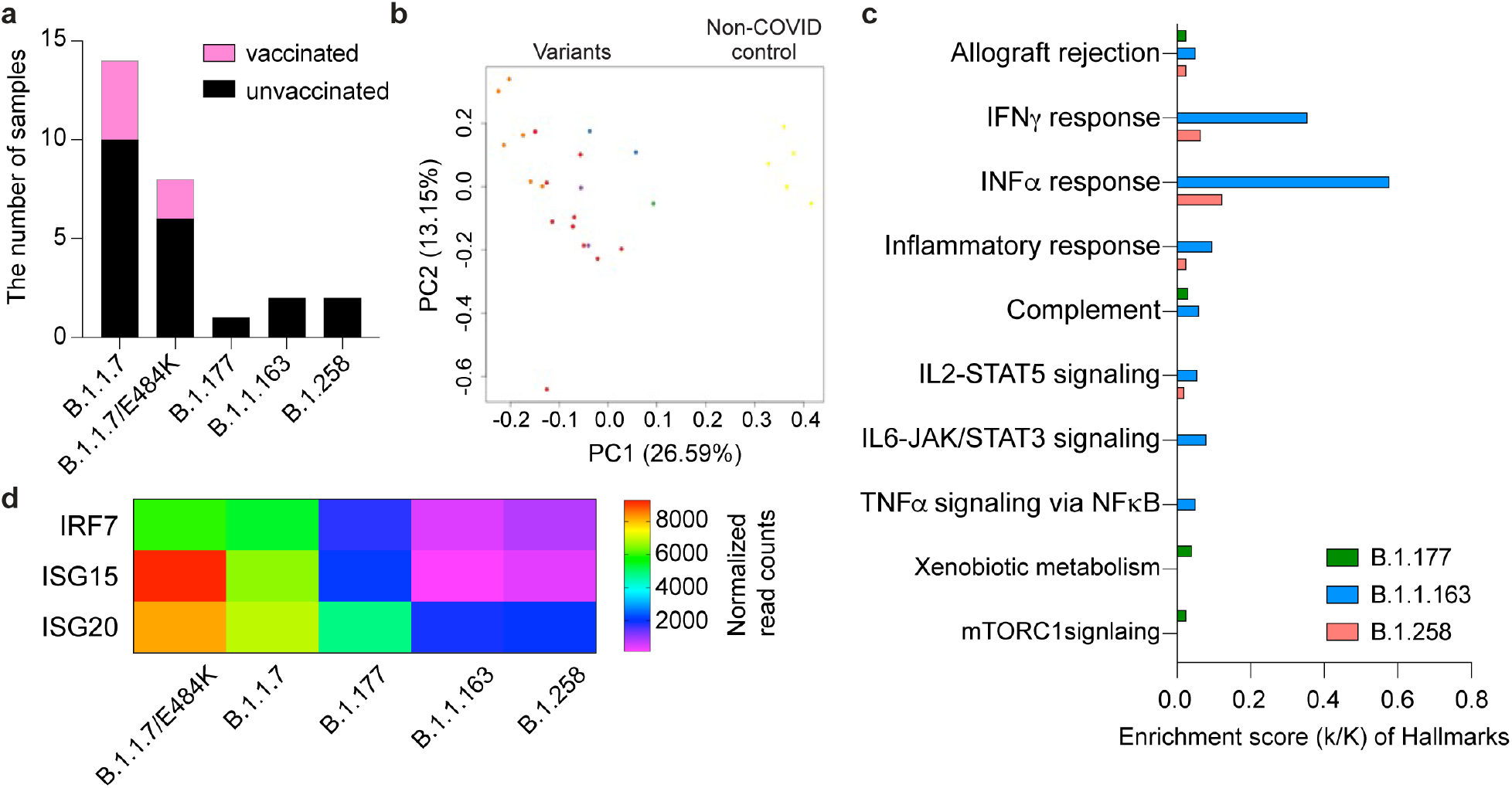
Differences of immune response between COVID-19 patients infected by different SARS-CoV-2 variants. **a**. The number of COVID-19 patients infected by each variant and vaccinated patient. **b**. Sparse PCA depicting global transcriptional profiles of unvaccinated and variant infected COVID-19 patients. Dark Brown: Non-COVID, Light Brown: E484K, Turquoise: B.1.1.7, Green: B.1.351, Orange: B.1.1.163, Purple: B.1.177, Red: B.1.258. **c**. Genes expressed at significantly higher levels in the B.1.1.7 infected COVID-19 patients compared to other variant infected patients were significantly enriched in immune regulation hallmarks (FDR q value < 0.005). **d**. The mRNA level of three significant genes in COVID-19 disease were presented using the heatmap.

## Discussion

As the COVID-19 epidemic has evolved, the impact of variants on disease progression and response to vaccine has become a key area of investigation^2,3^. The B.1.1.7 variant, first detected in Kent (UK) in September 2020, has spread rapidly and has become the predominant form in many countries^7,8^. The RBD mutation, E484K, first detected in the B.1.351 variant, has evolved independently in infected individuals in different genetic SARS-CoV-2 backgrounds. Here we were able to explore the impact of this escape mutation on the background of the B.1.1.7 variant^1,9^, utilizing samples from cohorts of hospitalized patients with the same demographics that resided in specific geographic regions and developed infection within the same timeframe, enabling a relatively controlled focus on the impact of this single amino acid substitution.

The clinical course of SARS-CoV-2 infection has been well-characterized, ranging from asymptomatic to fatal disease^10^. Here we concentrated on the immune transcriptome response of cohorts of more severely ill hospitalized patients and followed response over three weeks from the time of symptom development. In these patients we found that the host transcriptome response to SARS-CoV-2 infection evolves rapidly with the most profound changes within five days of symptomology, dropping significantly over the next ten days, with further drop, but not normalization, within four weeks. In a previous study we documented that asymptomatic and minimally symptomatic SARS-CoV-2 infected individuals from the same general geographic area showed no significant differences to uninfected individuals by six weeks following infection^6^. This means that studies of transcriptome responses following SARS-CoV-2 infection need to be controlled for time of infection and symptom development for valid comparisons. There was no significant difference in deaths between B.1.1.7 and B.1.1.7+E484K infection. Five and three individuals died in B.1.1.7 and B.1.1.7+E484K groups, respectively. Our study provided the opportunity to examine whether or not transcriptomes from those that survived were significantly different than those that expired from infection. No differences were found. All individuals who died were over the age of 70 and had underlying medical conditions. Drug treatment including corticosteroid and antibody treatment could theoretically alter immune response. In this study the majority of this hospitalized population received corticosteroid treatment, which may have modified immune response, but as there were no significant differences in the percentage treated between B.1.1.7 and B.1.1.7+E484K patients, this was not the reason for the significant differences in transcriptome response found. Only one individual within the cohort received therapy with convalescent plasma before the first blood sampling. Interestingly, the PCA demonstrated that this individual’s transcriptome mapped to a location different than the others. Due to this being a single sample, we cannot make any generalizations from this study about how antibody treatment may modify transcriptome response; however, this is an area of interest for future investigation if sufficient numbers of temporally and variant matched samples are available.

Prior vaccination, even a single dose, significantly modified transcriptome response, even as it did not prevent hospitalization. However, the significant differences in transcriptome response between B.1.1.7 and B.1.1.7+E484K infected individuals persisted even if these vaccinated individuals, documenting the reproducibility of the finding across clinical settings. Infection following first vaccination is well documented with some individuals also becoming severely ill after full vaccination^11^. Our study also included one individual who was hospitalized with infection following full vaccination but there were no significant differences in transcriptome response with the other vaccinated patients. Antibody titers were not available for any of the vaccinated individuals, but they did not fall into any of the currently recognized groups with inadequate immune response following vaccine.

A small number of samples from individuals infected with (B.1.351) and without (B.1.177, B.1.1.163, B.1.258) the E484K mutation (B.1.351) were available for comparison. Cohort sizes were not sufficient for statistical analyses of significance but temporally-matched transcriptomes did show variation from B.1.1.7 and B.1.1.7+E484K. A recent bulk RNA-seq immune transcriptome study also identified transcript categories, including interferon-activated genes, induced in COVID-19 patients but the variant was not defined^12^. As samples become available from patients with other variants, the degree of transcriptome variation will be able to be defined.

In summary, our study documents that the E484K mutation is sufficient to alter transcriptome response. This RBD escape mutation has evolved independently in several SARS-CoV-2 backgrounds, including B.1.1.7^9^ with a single immunocompromised patient study demonstrating that it can occur within 10 days of infection^13^. Here, our significant differences in transcriptome response did not correlate with significant differences in clinical course but this could be due to the selection bias of only hospitalized patients being included in this study. The fact that variants can induce different host immune response has implications for the understanding of both innate and acquired immune response mechanisms. Further investigation of the reasons why these differences occur may help in design of vaccines and antibody treatments that may have more universal coverage of variant.

### Limitations

The findings in this report are subject to several limitations. Our study focused on hospitalized patients in a specific geographic area. Our study focused on elderly patients with limited comparison to younger individuals. We have investigated the effect of the BNT162b vaccine but not of any other vaccine.

## Methods

### SARS-CoV-2 virus sequencing

RNA was extracted from patient’s blood using a Maxwell RSC simply RNA Blood purification kit according to the manufacturer’s instructions (Promega, USA). Library preparation and sequencing was performed as described^14^. In short, cDNA was obtained by using reverse transcriptase with random priming. Following cDNA synthesis, primers based on sequences from the ARTICnetwork were used to generate 400 bp amplicons in two different PCR pools. After merging of pools and amplification, libraries were constructed using QIASeq FX DNA Library UDI Kit following the manufacturer’s instructions (Qiagen GmbH, North Rhine-Westphalia, Germany).

Sequencing was done with Illumina NextSeq® 500/550 using 149-bp paired-end reads with 10-bp indices (Illumina, California, USA). Obtained viral sequences were assembled using CLC Genomics Workbench v20.0.3 (Qiagen GmbH, North Rhine-Westphalia, Germany). SARS-CoV-2 isolate Wuhan-Hu-1 served as the reference genome (Accession NC_045512.2). SARS-CoV-2 variants were identified by uploading FASTA-files on freely accessible databases (http://cov-lineages.org/).

### Extraction of the buffy coat and purification of RNA

Whole blood was collected, and total RNA was extracted from the buffy coat and purified using the Maxwell RSC simply RNA Blood Kit (Promega) according to the manufacturer’s instructions. The concentration and quality of RNA were assessed by an Agilent Bioanalyzer 2100 (Agilent Technologies, CA).

### mRNA sequencing (mRNA-seq) and data analysis

The Poly-A containing mRNA was purified by poly-T oligo hybridization from 1 μg of total RNA and cDNA was synthesized using SuperScript III (Invitrogen, MA). Libraries for sequencing were prepared according to the manufacturer’s instructions with TruSeq Stranded mRNA Library Prep Kit (Illumina, CA, RS-20020595) and paired-end sequencing was done with a NovaSeq 6000 instrument (Illumina).

The raw data were subjected to QC analyses using the FastQC tool (version 0.11.9) (https://www.bioinformatics.babraham.ac.uk/projects/fastqc/). mRNA-seq read quality control was done using Trimmomatic^15^ (version 0.36) and STAR RNA-seq^16^ (version STAR 2.5.4a) using 150 bp paired-end mode was used to align the reads (hg19). HTSeq^17^ (version 0.9.1) was to retrieve the raw counts and subsequently, R (https://www.R-project.org/), Bioconductor^18^ and DESeq2^19^ were used to normalize the counts across samples. Additionally, the RUVSeq^20^ package was applied to remove confounding factors. The data were pre-filtered keeping only genes with at least ten reads in total. The visualization was done using dplyr (https://CRAN.R-project.org/package=dplyr) and ggplot2^21^. Genes were categorized as significant differentially expressed with an adjusted p-value (pAdj) below 0.05 and a fold change > 2 for up-regulated genes and a fold change of < −2 for down-regulated ones with no zero values in all data set and then conducted gene enrichment analysis (GSEA, https://www.gsea-msigdb.org/gsea/msigdb).

## Data Availability

The RNA-seq data of Non-COVID individual were obtained from GSE162562. The RNA-seq data of COVID-19 patients infected by B.1.1.7 or B.1.1.7/E484K variant will be uploaded in GEO before publishing the manuscript.

## Statistical analysis

For significance of each GSEA categoty, significantly regulated gene sets were evaluated with the Kolmogorov-Smirnov statistic. Demographic data were analyzed by Chi-square on GraphPad Prism software (version 9.0.0). All graphs were generated using GraphPad.

## Ethics approval

This study was approved by the Institutional Review Board (IRB) of the Office of Research Oversight / Regulatory Affairs, Medical University of Innsbruck, Austria (EC numbers: 1064/2021).

## Acknowledgments

Our gratitude goes to the patients who contributed to this study to advance our understanding of COVID-19. This work was supported by the Intramural Research Programs (IRPs) of National Institute of Diabetes and Digestive and Kidney Diseases (NIDDK) and utilized the computational resources of the NIH HPC Biowulf cluster (http://hpc.nih.gov). RNA-sequencing was conducted in the NIH Intramural Sequencing Center, NISC (https://www.nisc.nih.gov/contact.htm).

## Author contribution

LK: recruited patients, collected material, analyzed data; HKL: analyzed data, wrote manuscript; MW: collected and prepared material; AM, AZ, LK Sr, JS and NK: recruited and diagnosed patients; PAF: analyzed data, wrote manuscript; LH: analyzed data, wrote manuscript. All authors read and approved the manuscript

## References

1. England, P.H., https://assets.publishing.service.gov.uk/government/uploads/system/uploads/attachment_data/file/959426/Variant_of_Concern_VOC_202012_01_Technical_Briefing_5.pdf (2021).

2. Collier, D.A. et al. Sensitivity of SARS-CoV-2 B.1.1.7 to mRNA vaccine-elicited antibodies. Nature (2021).

3. Wu, K. et al. Serum Neutralizing Activity Elicited by mRNA-1273 Vaccine. N Engl J Med (2021).

4. Mwenda, M. et al. Detection of B.1.351 SARS-CoV-2 Variant Strain - Zambia, December 2020. MMWR Morb Mortal Wkly Rep 70, 280–282 (2021).

5. Garcia-Beltran, W.F. et al. Multiple SARS-CoV-2 variants escape neutralization by vaccine-induced humoral immunity. Cell 184, 2372–2383 e9 (2021).

6. Lee, H.K. et al. Immune transcriptomes of highly exposed SARS-CoV-2 asymptomatic seropositive versus seronegative individuals from the Ischgl community. Sci Rep 11, 4243 (2021).

7. Pango. https://cov-lineages.org/global_report_B.1.1.7.html.

8. Frampton, D. et al. Genomic characteristics and clinical effect of the emergent SARS-CoV-2 B.1.1.7 lineage in London, UK: a whole-genome sequencing and hospital-based cohort study. Lancet Infect Dis (2021).

9. Moustafa, A.M. et al. Comparative Analysis of Emerging B.1.1.7+E484K SARS-CoV-2 isolates from Pennsylvania. bioRxiv (2021).

10. Lu, T. & Reis, B.Y. Internet search patterns reveal clinical course of COVID-19 disease progression and pandemic spread across 32 countries. NPJ Digit Med 4, 22 (2021).

11. Knabl, L. et al. Impact of BNT162b first vaccination on the immune transcriptome of elderly patients infected with the B.1.351 SARS-CoV-2 variant. medRxiv (2021).

12. McClain, M.T. et al. Dysregulated transcriptional responses to SARS-CoV-2 in the periphery. Nat Commun 12, 1079 (2021).

13. Lohr, B., Niemann, D. & Verheyen, J. Bamlanivimab treatment leads to rapid selection of immune escape variant carrying E484K mutation in a B.1.1.7 infected and immunosuppressed patient. Clin Infect Dis (2021).

14. Itokawa, K., Sekizuka, T., Hashino, M., Tanaka, R. & Kuroda, M. Disentangling primer interactions improves SARS-CoV-2 genome sequencing by multiplex tiling PCR. PLoS One 15, e0239403 (2020).

15. Bolger, A.M., Lohse, M. & Usadel, B. Trimmomatic: a flexible trimmer for Illumina sequence data. Bioinformatics 30, 2114–20 (2014).

16. Dobin, A. et al. STAR: ultrafast universal RNA-seq aligner. Bioinformatics 29, 15–21 (2013).

17. Anders, S., Pyl, P.T. & Huber, W. HTSeq--a Python framework to work with high-throughput sequencing data. Bioinformatics 31, 166–9 (2015).

18. Huber, W. et al. Orchestrating high-throughput genomic analysis with Bioconductor. Nat Methods 12, 115–21 (2015).

19. Love, M.I., Huber, W. & Anders, S. Moderated estimation of fold change and dispersion for RNA-seq data with DESeq2. Genome Biol 15, 550 (2014).

20. Risso, D., Ngai, J., Speed, T.P. & Dudoit, S. Normalization of RNA-seq data using factor analysis of control genes or samples. Nat Biotechnol 32, 896–902 (2014).

21. Wickham, H. Ggplot2 : elegant graphics for data analysis, viii, 212 p. (Springer, New York, 2009).

